# Does ECG-Based AI Detect Aortic Stenosis Beyond Conventional LVH Criteria? An Analysis of the CLIDAS Database

**DOI:** 10.64898/2026.06.07.26355087

**Authors:** Takenobu Shimada, Satoshi Kodera, Shinnosuke Sawano, JingChuan Guan, Wataru Saitoh, Shiho Wakasa, Shinji Ito, Tomoya Yanagishita, Yusuke Hayashi, Atsushi Shibata, Asahiro Ito, Kenichiro Otsuka, Yasutomi Higashikuni, Hiroshi Okamura, Kenichi Tsujita, Koichi Node, Osamu Yamaguchi, Hisaki Makimoto, Tomoyuki Kabutoya, Yasushi Imai, Masaharu Nakayama, Hisahiko Sato, Hideo Fujita, Takahide Kohro, Tetsuya Matoba, Norihiko Takeda, Daiju Fukuda, Ryozo Nagai, the CLIDAS Research Group

## Abstract

**Background:** Aortic stenosis (AS) is a progressive valvular disease associated with poor prognosis once symptoms develop, yet routine echocardiographic screening is impractical. While artificial intelligence (AI)-based electrocardiogram (ECG) models have shown promise for AS detection, it remains unclear whether they primarily reflect conventional left ventricular hypertrophy (LVH) voltage criteria or capture additional ECG features.

**Methods and Results:** We developed a deep learning model using 244,816 ECGs from 51,713 patients across six academic institutions in Japan (CLIDAS database). AS labels were derived from inpatient Diagnosis Procedure Combination (DPC) codes. The model achieved an area under the receiver operating characteristic curve (AUC) of 0.849 (95% confidence interval 0.832–0.865) in the independent test cohort, with consistent performance across institutions, sex, and age. At a threshold of 0.1, sensitivity was 79.1%, specificity was 73.9%, and negative predictive value (NPV) was 98.0%. Conventional LVH voltage criteria (Sokolow–Lyon AUC 0.706; Cornell AUC 0.692) showed lower performance, and adding them to the AI model conferred no incremental benefit (AUC 0.849 vs. 0.847). Gradient-weighted class activation mapping (Grad-CAM) revealed predominant attention around QRS complexes in limb leads, beyond regions typically assessed in LVH evaluation.

**Conclusions:** This multicenter AI-ECG model demonstrated strong discrimination for AS and captured ECG features beyond conventional LVH voltage criteria. The high NPV supports its use as a rule-out pre-screening tool.

## Introduction

Aortic stenosis (AS) is one of the most common valvular heart diseases in elderly populations and is associated with a poor prognosis once symptoms develop^1^. Its prevalence rises sharply with age, affecting approximately 3–10% of individuals aged 75 years or older with moderate-to-severe AS^2–5^. Despite its clinical significance, a substantial proportion of patients remain undiagnosed until symptoms emerge or AS is identified incidentally^6^.

Early detection is essential, as it enables timely referral for echocardiographic evaluation, appropriate planning of transcatheter or surgical aortic valve replacement, and perioperative risk stratification. However, routine echocardiographic screening is impractical and cost-prohibitive at a population scale^7^. The 12-lead electrocardiogram (ECG), by contrast, is ubiquitous, inexpensive, and routinely acquired in a wide range of clinical settings, making it an attractive medium for opportunistic screening.

Recent advances in deep learning have enabled artificial intelligence (AI)-based ECG models to detect a variety of structural and functional cardiac conditions^8–14^. Several studies have reported promising discriminative performance for AS detection using AI-ECG models. However, a critical mechanistic question remains unresolved: whether these models merely recapitulate conventional electrocardiographic markers of left ventricular hypertrophy (LVH) or capture additional pathophysiological features beyond these criteria.

The present study had two aims: to develop a deep learning model for AS detection using a multicenter 12-lead ECG database, and to investigate whether the model’s predictions are primarily driven by conventional LVH voltage criteria or reflect broader ECG features of AS.

## Materials and methods

### CLIDAS database

The Clinical Deep Data Accumulation System (CLIDAS) database is a multicenter research platform developed to construct a large-scale real-world clinical data infrastructure across multiple academic institutions in Japan^15,16^. The database integrates digital 12-lead ECG recordings, chest radiographs, and comprehensive inpatient administrative data based on the Diagnosis Procedure Combination (DPC) system, a Japanese claims-based reimbursement system that records diagnoses for each hospitalization. By linking imaging data with patient-level DPC records, disease labels derived from real-world clinical coding can be assigned to ECG and radiographic data, enabling large-scale development of AI models. Of the participating institutions, six whose data were available at the time of this analysis contributed to the present study: Kyushu University, Saga University, the University of Tokyo, Ehime University, Osaka Metropolitan University, and Kumamoto University.

### Study design

This was a multicenter retrospective study using the CLIDAS database, covering the period from April 2013 to March 2024. All digital 12-lead ECGs recorded during this period at the participating institutions were screened for inclusion. ECGs were linked to corresponding DPC records within the CLIDAS database based on hospitalization dates. Recordings obtained within 28 days following the admission date were included, while ECGs recorded prior to admission were excluded, as pre-admission ECGs may not reflect the diagnosed condition. For patients with multiple hospitalizations, only the first admission was used for label assignment to avoid duplicate counting.

AS was defined as ICD-10 code I35.0 (non-rheumatic AS) recorded in any of the three DPC diagnostic fields (primary diagnosis, diagnosis triggering hospitalization, or diagnosis consuming the most medical resources).

The dataset was split at the patient level into training (70%), validation (15%), and test (15%) cohorts using a fixed random seed to ensure reproducibility, with patient-level splitting employed to prevent data leakage across cohorts (Figure 1). A deep learning model was developed using the training cohort to predict the probability of AS from 12-lead ECG signals. The optimal classification threshold was selected based on performance in the validation cohort. Final model performance was evaluated in the test cohort, which was not used for model development or threshold selection.

**Figure 1.**
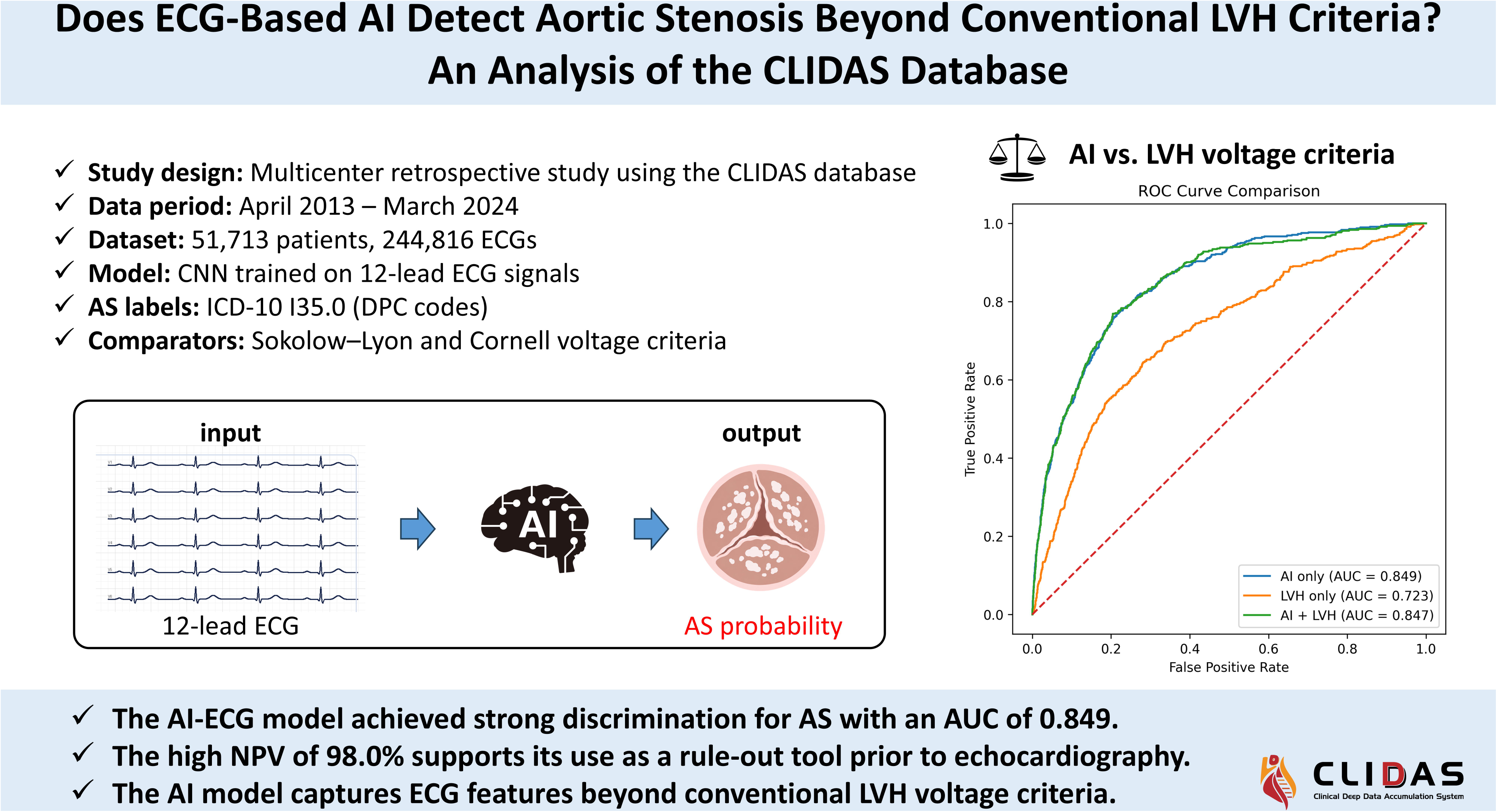
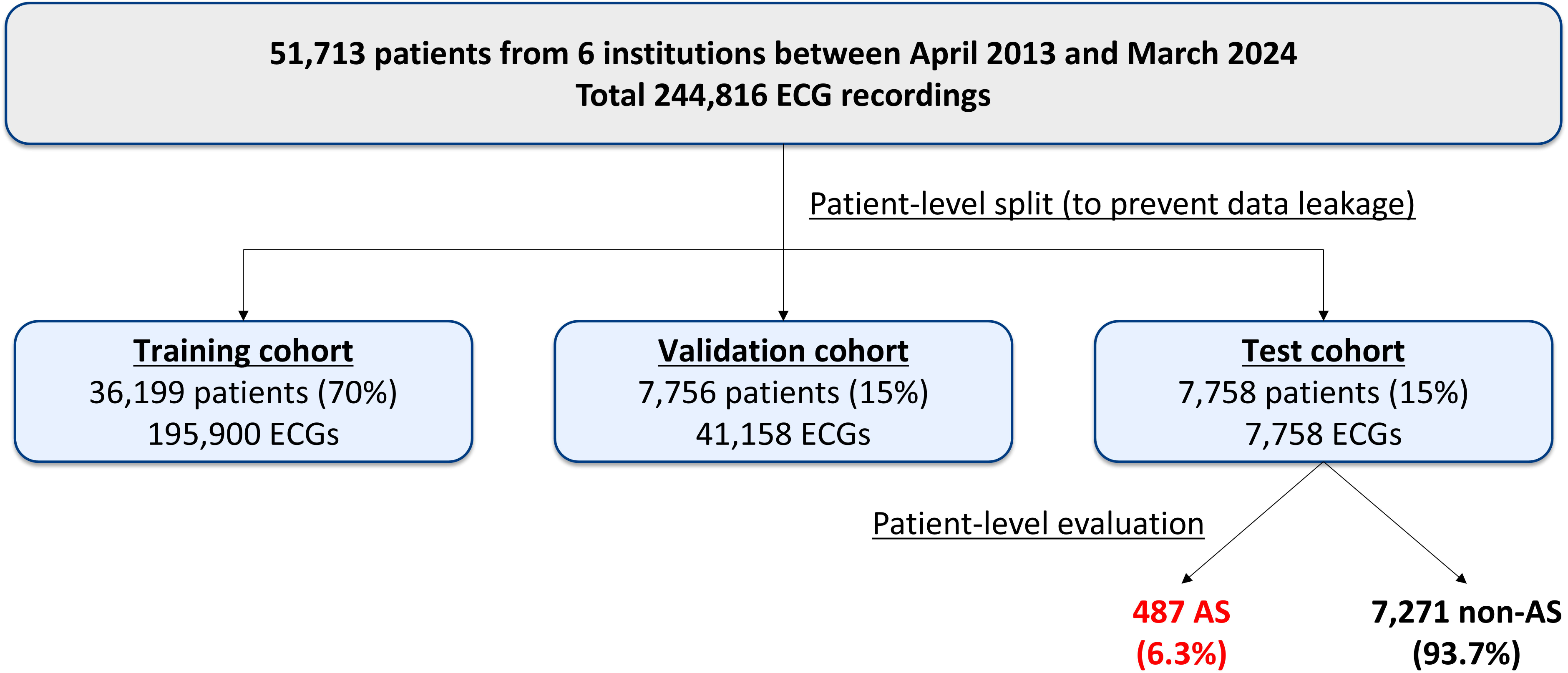
Study flow and cohort allocation Flowchart illustrating the study population and cohort allocation from six institutions between April 2013 and March 2024. Abbreviations: AS, aortic stenosis

To assess the generalizability of the model, performance was evaluated across institutions, as well as in subgroups defined by sex and age. To investigate whether the model’s predictions were driven by conventional LVH markers, we examined correlations between AI-derived prediction scores and two established LVH voltage criteria (Sokolow–Lyon and Cornell) and compared discriminative performance of the AI model with that of the LVH criteria alone and in combination. Gradient-weighted class activation mapping (Grad-CAM) was additionally applied to identify the ECG regions most influential in the model’s predictions.

This study complied with the Declaration of Helsinki for investigation in humans and was conducted with the approval of the Jichi Medical University Bioethics Committee for Medical Research (24-164). The requirement for written informed consent was waived owing to the retrospective design of the study, and an opt-out approach was employed to inform participants. The study is registered with the University Hospital Medical Information Network Clinical Trial Registry (UMIN000057338).

### ECG preprocessing

Raw 12-lead ECG signals were converted to mV according to institution-specific file formats and standardized to 5,000 samples per recording. Recordings shorter than 5,000 samples were zero-padded at a random location, whereas longer recordings were randomly cropped to 5,000 samples. The sampling rate was inferred from the signal length and was either 500 Hz (corresponding to 10-second recordings) or 1,000 Hz (corresponding to 5-second recordings). Each lead was then normalized using the per-lead mean and standard deviation derived from the University of Tokyo ECG dataset.

### Model architecture

We developed a two-dimensional convolutional neural network (CNN) to predict the probability of AS from standard 12-lead ECG signals. Each input ECG was represented as a tensor of shape (12 leads × 5,000 samples) and treated as a two-dimensional input (lead × time).

The network comprised six temporal convolutional blocks, each consisting of 2D convolutions with kernels spanning the temporal dimension, batch normalization, ReLU activation, and max-pooling. The number of filters increased progressively from 16 in the first two blocks to 32 and 64 in subsequent blocks. Following these temporal blocks, a lead integration layer (a 2D convolution with a kernel spanning all 12 leads) to aggregate inter-lead information into a compact feature representation. This was passed through two fully connected layers (each with 256 units), with batch normalization and dropout (rate = 0.5) applied after each. The final output was a scalar probability of AS produced by a sigmoid activation. The model architecture is summarized in Supplemental Figure 1.

The model was trained using binary cross-entropy loss with the Adam optimizer (batch size = 128). The learning rate was selected from {1×10□□, 1×10□□, 5×10□□} based on validation AUC and was reduced by a factor of 0.5 if no improvement was observed over three consecutive epochs. Training was proceeded for up to 30 epochs, with early stopping after 15 epochs without improvement in validation AUC.

### Threshold selection

The optimal classification threshold was determined using the validation cohort to prevent data leakage from the test cohort. Candidate thresholds of 0.05, 0.10, 0.20, and 0.50 were evaluated based on the balance between sensitivity and specificity. The threshold that provided the most clinically reasonable balance, prioritizing sensitivity for screening purposes, was selected and subsequently applied to the test cohort for all performance evaluations.

### Conventional LVH voltage criteria

Two widely used voltage-based ECG criteria for LVH were calculated from the raw ECG waveforms as comparators for the AI model predictions. The Sokolow–Lyon voltage was defined as the sum of the S-wave amplitude in lead V1 and the R-wave amplitude in lead V5 or V6^17^. The Cornell voltage was defined as the sum of the R-wave amplitude in lead aVL and the S-wave amplitude in lead V3^18^. Conventional thresholds for LVH (e.g., ≥3.5 mV for Sokolow–Lyon and sex-specific thresholds for Cornell voltage) were provided for reference. In the present study, these voltage values were analyzed as continuous variables.

Voltage amplitudes were extracted from the raw ECG signals using an automated algorithm. The R-peak was identified using lead II, and S-wave amplitude was defined as the minimum value within an 80 ms window following each R-peak (Supplemental Figure 2). Values were aggregated across beats using the median. Recordings with voltage values exceeding 20 mV in absolute value were excluded as physiologically implausible artifacts (n = 2).

### Statistical analysis

The primary performance metric was the area under the receiver operating characteristic curve (AUC). The 95% confidence intervals (CIs) for AUC was estimated by patient-level bootstrapping with 2,000 resamples (random seed = 42). Subgroup AUC analyses were performed by institution, sex (male / female), and age group (<80 years / ≥80 years).

At the pre-specified threshold selected from the validation cohort, screening performance in the test cohort was evaluated by calculating sensitivity, specificity, positive predictive value (PPV), and negative predictive value (NPV) with 95% CIs.

The relationship between AI prediction scores and LVH voltage indices was assessed using Spearman’s rank correlation coefficient. The discriminative performance of the AI model was compared with that of the Sokolow–Lyon voltage criterion alone, the Cornell voltage criterion alone, and a logistic regression model combining both voltage criteria, using AUC as the primary metric. AUC comparisons were performed using DeLong’s method. The incremental value of adding LVH voltage indices to the AI model was evaluated by comparing the AUC of the AI model alone with that of a logistic regression model incorporating both the AI prediction score and LVH voltage indices.

Grad-CAM (gradient-weighted class activation mapping) was applied to representative true-positive and false-negative cases to identify ECG regions with high model attention.

All statistical analyses were performed using Python (version 3.10.12). Statistical significance was defined as p<0.05 (two-sided).

## Results

### Study population

A total of 244,816 ECG recordings from 51,713 unique patients, linked to DPC records, were included in the final dataset (training: 195,900 ECG recordings from 36,199 patients; validation: 41,158 ECG recordings from 7,756 patients; test: 7,758 ECG recordings, one per patient, from 7,758 patients). The mean age was 67.4 ± 14.7 years and 32,035 patients (61.9%) were male. Detailed patient characteristics are summarized in Table 1.

The proportion of AS-positive patients was 6.6% in the training cohort, 6.7% in the validation cohort, and 6.3% in the test cohort.

### Model performance in the test cohort

In the test cohort, the AI model achieved strong discrimination with an AUC of 0.849 (95% CI, 0.832–0.865) (Figure 2), with consistent performance across institutions, sex, and age groups (Supplemental Table 1). A classification threshold of 0.1 was selected from the validation cohort, prioritizing sensitivity for screening purposes (Supplemental Table 2). At this threshold, the model demonstrated a sensitivity of 79.1% and specificity of 73.9%, with a PPV of 18.0% and NPV of 98.0% in the test cohort (Figure 3).

**Figure 2.**
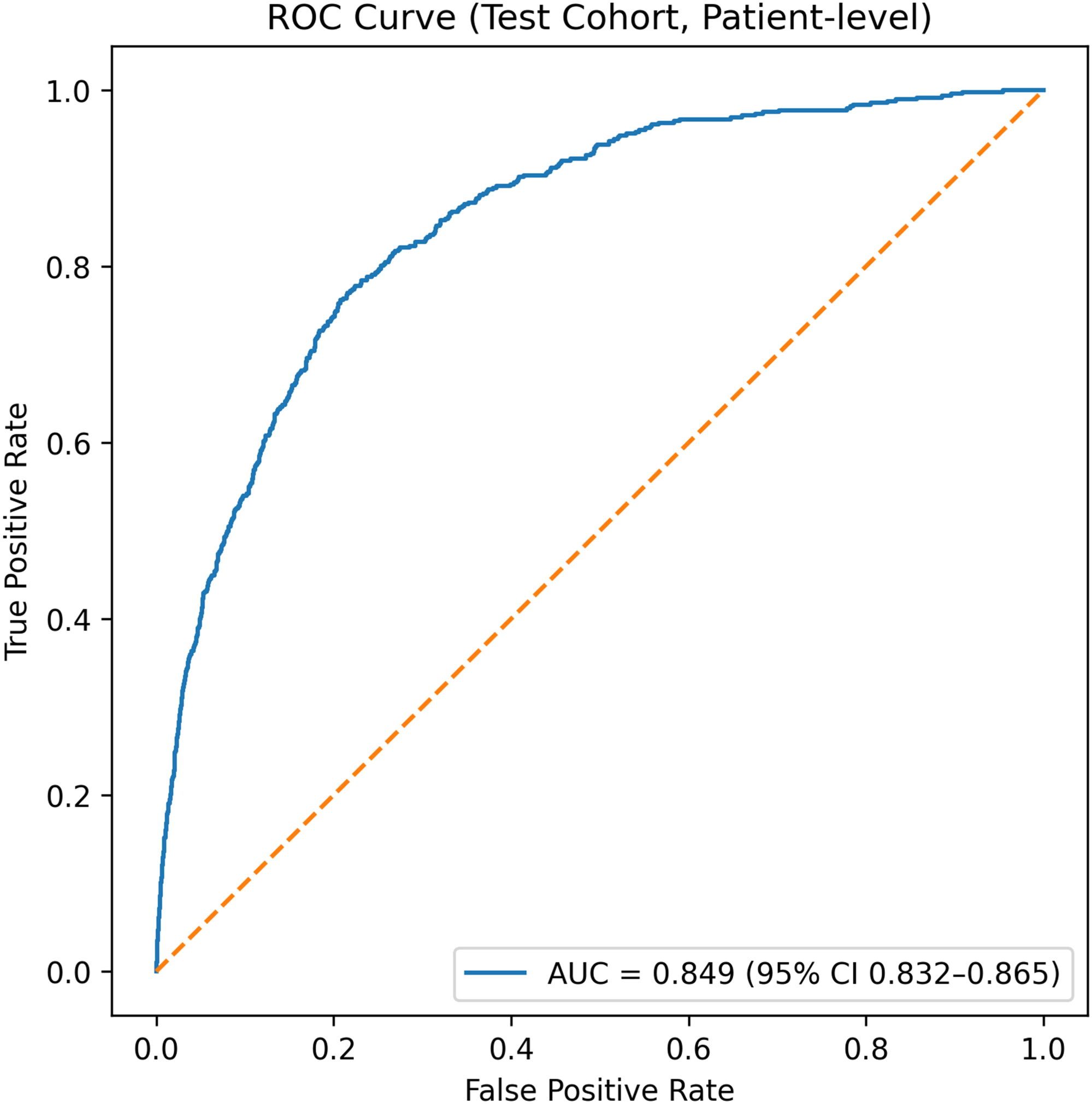
Receiver operating characteristic curve for AI-ECG model performance in the test cohort The deep learning model demonstrated strong discriminative performance in the independent test cohort (n = 7,758), achieving an AUC of 0.849 (95% CI 0.832–0.865). Abbreviations: AI, artificial intelligence; ECG, electrocardiogram; ROC, receiver operating characteristic; AUC, area under the curve; CI, confidence interval

**Figure 3.**
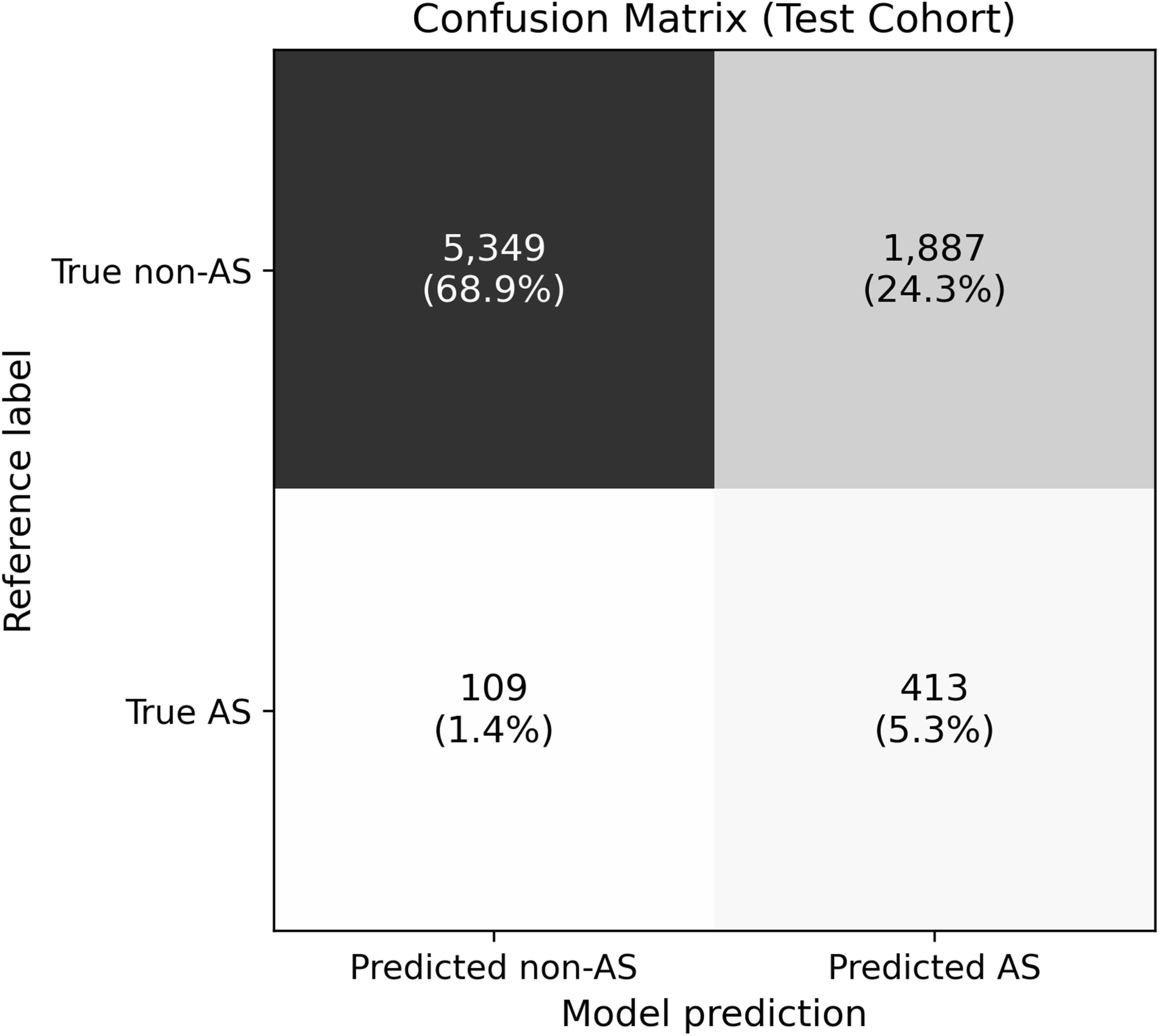
Confusion matrix of AI-ECG model predictions at a probability threshold of 0.1 Confusion matrix showing the classification performance of the AI model in the test cohort (n = 7,758) at a prediction probability threshold of 0.1. Numbers represent patient counts and percentages. Abbreviations: AI, artificial intelligence; ECG, electrocardiogram; AS, aortic stenosis

### Comparison with conventional LVH voltage criteria

The AI prediction score showed a modest positive correlation with both the Sokolow–Lyon and Cornell voltage (Spearman’s ρ = 0.385 and 0.329, respectively); however, there was substantial overlap between AS and non-AS groups (Figure 4). The AUCs of the Sokolow–Lyon criterion and Cornell voltage criterion alone for predicting AS were 0.706 and 0.692, respectively (Supplemental Figure 3). Figure 5 shows the ROC curves comparing AI alone, LVH indices alone, and AI combined with LVH indices. The curve for AI plus LVH indices almost completely overlaps with that for AI alone. Adding LVH voltage criteria to the AI model did not meaningfully improve discriminative performance (AUC 0.849 vs. 0.847 for AI alone vs. AI + LVH combined), suggesting that the AI model not only captures LVH-related ECG features, but also incorporates additional features beyond conventional voltage criteria.

**Figure 4.**
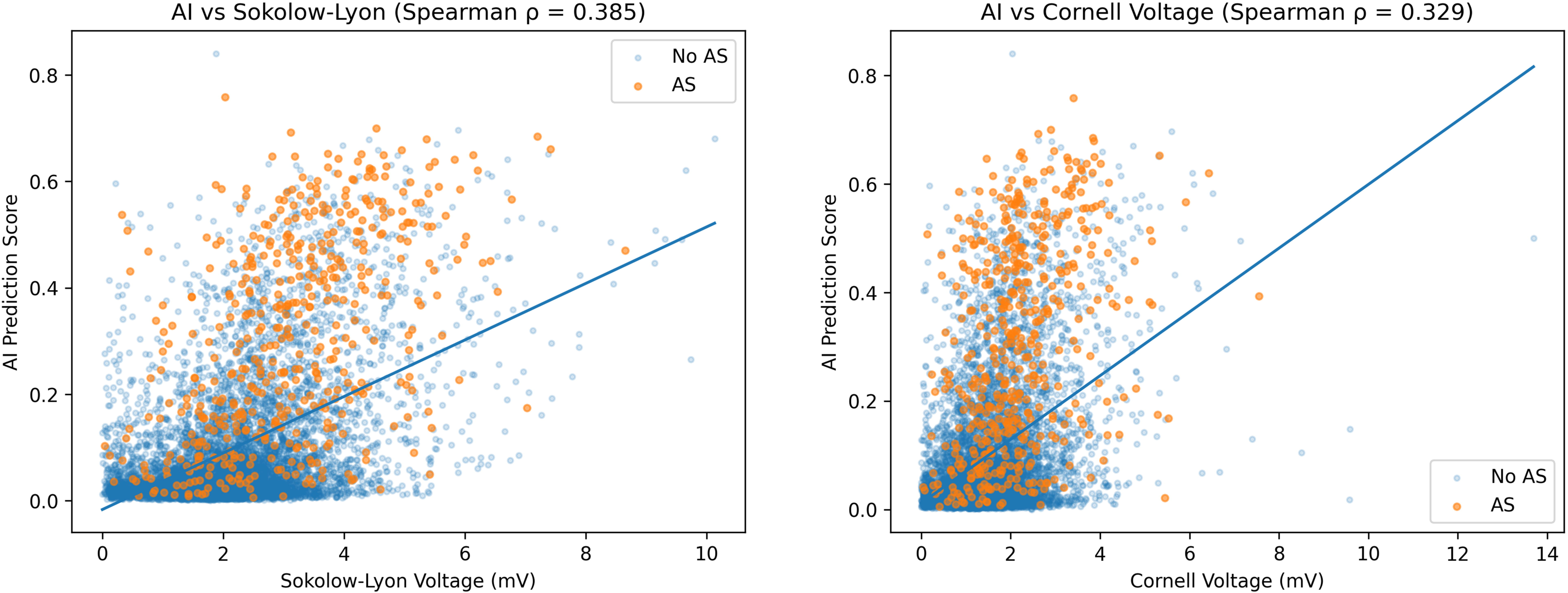
Correlation between AI prediction scores and conventional LVH voltage indices Scatter plots showing the relationship between AI prediction scores and Sokolow–Lyon voltage (left, Spearman’s ρ = 0.385) and Cornell voltage (right, Spearman’s ρ = 0.329) in the test cohort. Each point represents one patient, colored by AS status (orange: AS, blue: non-AS). Regression lines are shown in blue. Despite statistically significant correlations, substantial overlap between AS and non-AS groups was observed across all voltage ranges. Abbreviations: AI, artificial intelligence; LVH, left ventricular hypertrophy; AS, aortic stenosis; ECG, electrocardiogram

**Figure 5.**
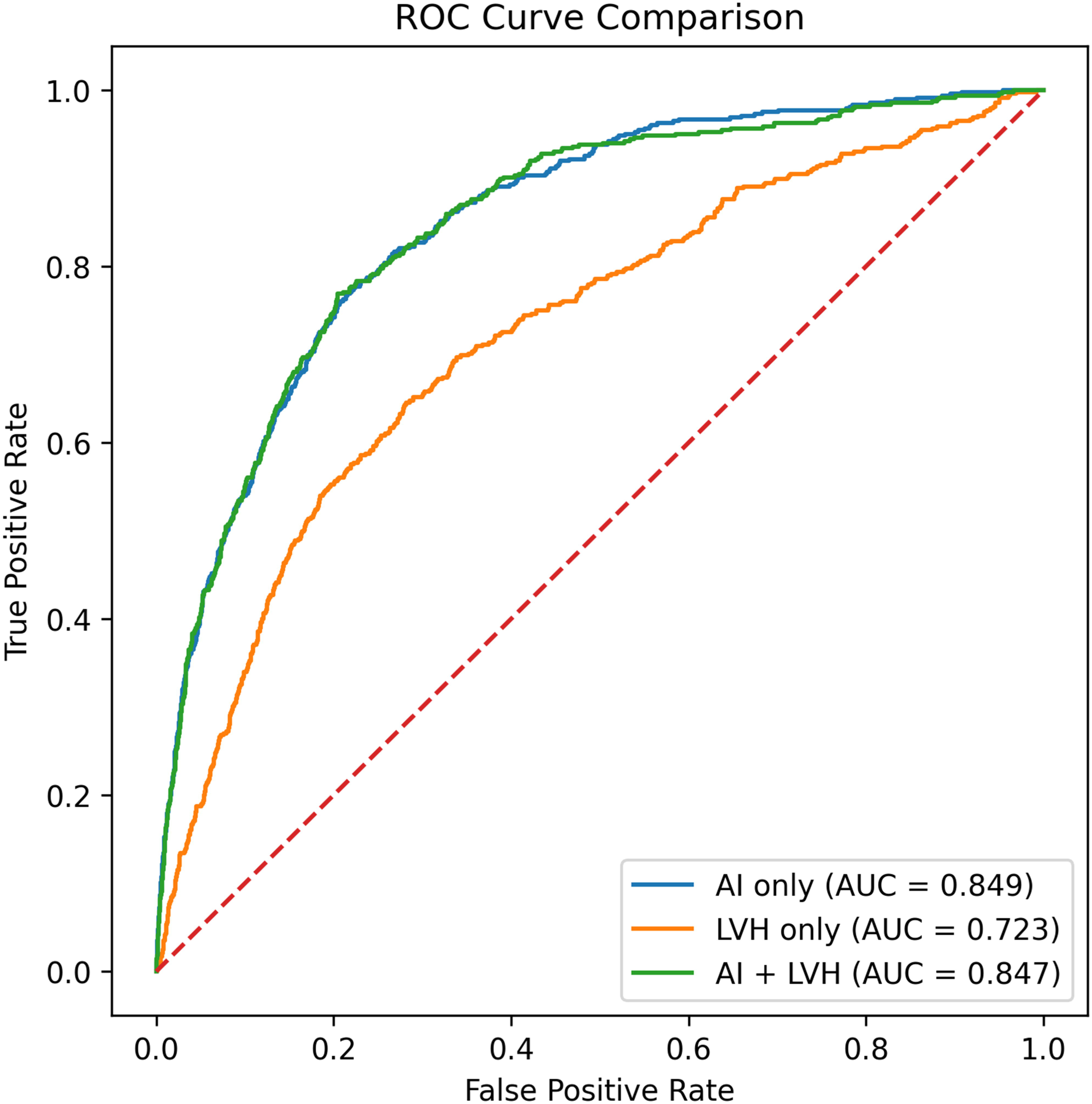
Comparison of discriminative performance: AI model alone versus LVH voltage criteria alone versus combined model ROC curves comparing three predictive models in the test cohort: the AI-ECG model alone (AUC = 0.849), combined Sokolow–Lyon and Cornell voltage criteria alone (AUC = 0.723), and the combined model incorporating both (AUC = 0.847). The addition of LVH voltage criteria to the AI model did not meaningfully improve discriminative performance. Abbreviations: AI, artificial intelligence; ECG, electrocardiogram; LVH, left ventricular hypertrophy; AUC, area under the curve; ROC, receiver operating characteristic

### Grad-CAM analysis

Grad-CAM analysis suggested that the model primarily focused on QRS complexes, with notable attention in limb leads (Figure 6). These patterns were not restricted to the precordial leads and voltage amplitudes typically emphasized in conventional LVH assessment.

**Figure 6.**
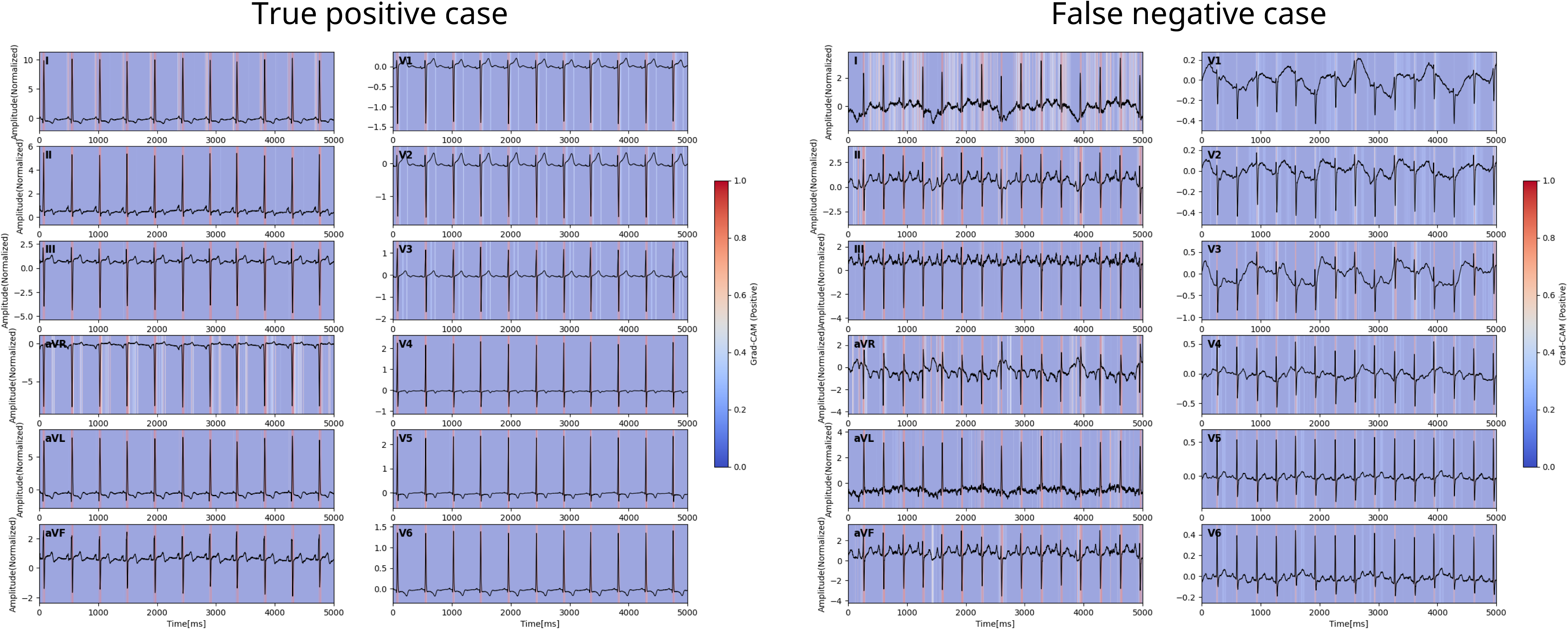
Representative Grad-CAM activation maps in true positive and false negative cases Grad-CAM heatmaps overlaid on 12-lead ECG recordings from a representative true positive case (left, correctly identified AS) and a false negative case (right, AS missed by the model). Warmer colors (red) indicate regions of greater model attention, while cooler colors (blue) indicate lower attention. Abbreviations: Grad-CAM, gradient-weighted class activation mapping; ECG, electrocardiogram; AS, aortic stenosis

## Discussion

The main findings of this study were threefold. First, the AI-ECG model achieved strong discrimination for AS across six academic institutions (AUC 0.849), with consistent performance across institutions, sex and age subgroups. Second, at a screening threshold of 0.1, the model achieved a high NPV of 98.0%, supporting its use as a rule-out tool prior to echocardiography. Third, conventional LVH voltage criteria alone showed markedly lower discriminative performance, and adding them to the AI model conferred no incremental benefit, indicating that the AI model captures ECG features beyond voltage criteria.

While prior studies have reported AI-ECG models for AS detection, few studies have systematically quantified whether model performance exceeds that of conventional LVH voltage criteria. This study represents one of the largest multicenter AI-ECG studies of AS in an Asian population, and among the first to systematically quantify the incremental value beyond conventional LVH voltage criteria.

AI prediction scores showed a moderate but statistically significant correlation with Sokolow–Lyon and Cornell voltage criteria, confirming that the model partially incorporates LVH-related voltage information. However, the substantially higher discriminative performance of the AI model and the absence of incremental benefit from adding LVH indices to the AI model together indicate that the model extracts substantial additional signal from the ECG waveform.

Grad-CAM analysis revealed that the model’s attention was predominantly concentrated around the QRS complex, with notable contributions from limb leads, a pattern not restricted to the precordial leads typically emphasized in visual LVH assessment. This QRS-centric attention may reflect morphological features of LV pressure overload beyond simple voltage amplitude, such as QRS duration, axis deviation, and subtle waveform changes that conventional voltage criteria do not capture.

In this study, AS labels were derived from inpatient DPC diagnosis codes rather than echocardiographic confirmation, a pragmatic choice that enabled large-scale multicenter data assembly. Potential concerns include coding inaccuracy and the fact that ICD-10 code I35.0 does not distinguish AS severity. Nonetheless, given that DPC codes are assigned to inpatients, the study population likely reflects clinically relevant AS rather than incidentally detected mild disease. Future work linking ECG data to echocardiographic severity measures would allow more precise clinical positioning.

The high NPV of 98.0% at a threshold of 0.1 represents a key clinical implication of this study. In practical terms, a negative AI-ECG result may effectively rule out AS in the screened population, supporting its use as a pre-screening tool prior to echocardiography. This rule-out utility is most applicable in settings where the ECG is already routinely performed but echocardiographic capacity is limited, such as opportunistic screening of elderly patients in primary care, pre-operative assessment, and routine health check-ups.

From a clinical perspective, auscultation remains the foundation of initial AS detection, as a systolic ejection murmur is a key finding. The AI-ECG model is best conceptualized not as a replacement for clinical evaluation, but as a complementary tool to support decision-making in settings where auscultation findings are equivocal or physical examination is limited, potentially prompting echocardiographic referral in patients who might otherwise be overlooked.

Future studies linking AI-ECG predictions to echocardiographic severity data will be important to clarify which disease stages the model most effectively identifies. Furthermore, longitudinal tracking of AI-ECG scores in individual patients may offer a non-invasive means of monitoring AS progression and informing the timing of echocardiographic follow-up, and represents a promising direction for future research.

## Limitations

Several limitations of this study should be acknowledged. First, AS labels were derived from inpatient DPC diagnosis codes (ICD-10 code I35.0) rather than echocardiographic confirmation. This code encompasses a broad spectrum of AS severity, and the model was therefore trained on the binary outcome of any AS versus no AS without severity stratification. Although this approach enabled large-scale multicenter data assembly, coding inaccuracies and incomplete capture of all AS patients cannot be excluded. Second, the study population was derived exclusively from inpatients at academic institutions in Japan. This population may not be representative of community-dwelling populations or primary care settings, in which the prevalence and clinical characteristics of AS may differ substantially. Third, although performance was consistent across six institutions, all sites were part of the same research project, and validation in fully independent external cohorts remains an important next step. Fourth, Grad-CAM was applied to representative cases for illustrative purposes, and the identified regions of model attention should be interpreted with caution as a qualitative indication of model attention rather than a mechanistic explanation of the pathophysiological features learned.

## Conclusion

This multicenter AI-ECG model achieved strong discrimination for AS and captured ECG features beyond conventional LVH voltage criteria. The high NPV supports its use as a rule-out pre-screening tool to complement routine clinical assessment.

## Acknowledgements

This work was supported by the Cross-ministerial Strategic Innovation Promotion Program (SIP) on “Integrated Health Care System” Grant Number JPJ012425. The authors thank all members of the CLIDAS Research Group for their contributions.

## Data availability

The data that support the findings of this study are available from the corresponding author upon reasonable request, subject to approval by the CLIDAS Research Group.

## Notes

### Competing Interest Statement

Satoshi Kodera and Norihiko Takeda have joint research agreements with Fukuda Denshi. All other authors have no conflicts of interest relevant to this study to declare.

### Author Declarations

This study was approved by the Jichi Medical University Bioethics Committee for Medical Research (24-164).

## References

1. Ross J, Braunwald E. Aortic Stenosis. Circulation 1968;38. doi:10.1161/01.CIR.38.1S5.V-61.

2. Osnabrugge RLJ, Mylotte D, Head SJ, Van Mieghem NM, Nkomo VT, LeReun CM, et al. Aortic Stenosis in the Elderly. J Am Coll Cardiol 2013;62:1002–1012. doi:10.1016/j.jacc.2013.05.015.

3. Lindroos M, Kupari M, Heikkilä J, Tilvis R. Prevalence of aortic valve abnormalities in the elderly: An echocardiographic study of a random population sample. J Am Coll Cardiol 1993;21:1220–1225. doi:10.1016/0735-1097(93)90249-Z.

4. Osnabrugge RLJ, Mylotte D, Head SJ, Van Mieghem NM, Nkomo VT, LeReun CM, et al. Aortic Stenosis in the Elderly. J Am Coll Cardiol 2013;62:1002–1012. doi:10.1016/j.jacc.2013.05.015.

5. Eveborn GW, Schirmer H, Heggelund G, Lunde P, Rasmussen K. The evolving epidemiology of valvular aortic stenosis. The Tromsø Study. Heart 2013;99:396–400. doi:10.1136/heartjnl-2012-302265.

6. d’Arcy JL, Coffey S, Loudon MA, Kennedy A, Pearson-Stuttard J, Birks J, et al. Large-scale community echocardiographic screening reveals a major burden of undiagnosed valvular heart disease in older people: the OxVALVE Population Cohort Study. Eur Heart J 2016;37:3515–3522. doi:10.1093/eurheartj/ehw229.

7. Papolos A, Narula J, Bavishi C, Chaudhry FA, Sengupta PP. U.S. Hospital Use of Echocardiography. J Am Coll Cardiol 2016;67:502–511. doi:10.1016/j.jacc.2015.10.090.

8. Elias P, Poterucha TJ, Rajaram V, Moller LM, Rodriguez V, Bhave S, et al. Deep Learning Electrocardiographic Analysis for Detection of Left-Sided Valvular Heart Disease. J Am Coll Cardiol 2022;80:613–626. doi:10.1016/j.jacc.2022.05.029.

9. Kwon J, Lee SY, Jeon K, Lee Y, Kim K, Park J, et al. Deep Learning–Based Algorithm for Detecting Aortic Stenosis Using Electrocardiography. J Am Heart Assoc 2020;9. doi:10.1161/JAHA.119.014717.

10. Cohen-Shelly M, Attia ZI, Friedman PA, Ito S, Essayagh BA, Ko W-Y, et al. Electrocardiogram screening for aortic valve stenosis using artificial intelligence. Eur Heart J 2021;42:2885–2896. doi:10.1093/eurheartj/ehab153.

11. Sawano S, Kodera S, Katsushika S, Nakamoto M, Ninomiya K, Shinohara H, et al. Deep learning model to detect significant aortic regurgitation using electrocardiography. J Cardiol 2022;79:334–341. doi:10.1016/j.jjcc.2021.08.029.

12. Sato M, Kodera S, Setoguchi N, Tanabe K, Kushida S, Kanda J, et al. Deep Learning Models for Predicting Left Heart Abnormalities From Single-Lead Electrocardiogram for the Development of Wearable Devices. Circulation Journal 2023;88:CJ-23-0216. doi:10.1253/circj.CJ-23-0216.

13. Kishikawa R, Kodera S, Setoguchi N, Tanabe K, Kushida S, Nanasato M, et al. An ensemble learning model for detection of pulmonary hypertension using electrocardiogram, chest X-ray, and brain natriuretic peptide. European Heart Journal - Digital Health 2025;6:209–217. doi:10.1093/ehjdh/ztae097.

14. Fujiki G, Kodera S, Setoguchi N, Tanabe K, Miyaji K, Kushida S, et al. Deep Learning-Based Identification of Echocardiographic Abnormalities From Electrocardiograms. JACC: Asia 2025;5:88–98. doi:10.1016/j.jacasi.2024.10.012.

15. Matoba T, Kohro T, Fujita H, Nakayama M, Kiyosue A, Miyamoto Y, et al. Architecture of the Japan Ischemic Heart Disease Multimodal Prospective Data Acquisition for Precision Treatment (J-IMPACT) System. Int Heart J 2019;60:264–270. doi:10.1536/ihj.18-113.

16. Matoba T, Katsuki S, Nakano Y, Kawahara T, Kimura M, Hino R, et al. Efficacy and Safety of High-Intensity Statins in Japanese Patients After Percutaneous Coronary Intervention ― Insights From the Clinical Deep Data Accumulation System (CLIDAS&lt;sup&gt;®&lt;/sup&gt;) ―. Circulation Journal 2025;89:CJ-25-0066. doi:10.1253/circj.CJ-25-0066.

17. Sokolow M, Lyon TP. The ventricular complex in left ventricular hypertrophy as obtained by unipolar precordial and limb leads. Am Heart J 1949;37:161–186. doi:10.1016/0002-8703(49)90562-1.

18. Casale PN, Devereux RB, Kligfield P, Eisenberg RR, Miller DH, Chaudhary BS, et al. Electrocardiographic detection of left ventricular hypertrophy: Development and prospective validation of improved criteria. J Am Coll Cardiol 1985;6:572–580. doi:10.1016/S0735-1097(85)80115-7.

