## Supplementary material for "Does ECG-Based AI Detect Aortic Stenosis Beyond Conventional LVH Criteria? An Analysis of the CLIDAS Database": Table 1

**Table 1. Baseline characteristics of the entire study population**

|  | n = 51,713 |
| --- | --- |
| Age, y | 67.4±14.7 |
| Male, n (%) | 32,035 (61.9) |
| Hypertension, n (%) | 19,386 (37.5) |
| Diabetes mellitus, n (%) | 10,670 (20.6) |
| Dyslipidemia, n (%) | 13,078 (25.3) |
| Renal disease, n (%) | 3,843 (7.4) |
| Chronic heart failure, n (%) | 18,151 (35.1) |
| Coronary artery disease, n (%) | 9,676 (18.7) |
| Atrial fibrillation, n (%) | 4,923 (9.5) |

Baseline characteristics of all patients included in the study (n = 51,713). Continuous variables are expressed as mean ± standard deviation, and categorical variables as number and percentage.

Abbreviations: y, years
