## Supplemental Appendix for "Does ECG-Based AI Detect Aortic Stenosis Beyond Conventional LVH Criteria? An Analysis of the CLIDAS Database"

| Table of contents | Page |
| --- | --- |
| CLIDAS Participating centers and investigators | 2 |
| Supplemental Figure 1 | 3 |
| Supplemental Figure 2 | 4 |
| Supplemental Table 1 | 5 |
| Supplemental Table 2 | 6 |
| Supplemental Figure 3 | 7 |

**CLIDAS Participating centers and investigators:**

Chiba University Graduate School of Medicine: Yoshio Kobayashi

Ehime University Graduate School of Medicine: Osamu Yamaguchi, Eizen Kimura

Jichi Medical University Saitama Medical Center: Hideo Fujita

Jichi Medical University School of Medicine: Ryozo Nagai, Takahide Kohro, Tomoyuki Kabutoya, Yasushi Imai, Hisaki Makimoto

Juntendo University Graduate School of Medicine: Tohru Minamino, Hiroshi Iwata

Kumamoto University: Kenichi Tsujita, Taishi Nakamura

Kyoto University Graduate School of Medicine: Koh Ono

Kyushu University Graduate School of Medical Sciences: Tetsuya Matoba, Yasuhiro Nakano

National Center for Global Health and Medicine: Yukio Hiroi, Kengo Miyo

National Cerebral and Cardiovascular Center: Takeshi Kitai

Osaka Metropolitan University Graduate School of Medicine: Daiju Fukuda

Osaka University Graduate School of Medicine: Yasushi Sakata, Toshihiro Takeda

Precision Inc.: Hisahiko Sato

Saga University: Koichi Node

The University of Tokyo Hospital: Satoshi Kodera, Norihiko Takeda

Tohoku University Graduate School of Medicine: Masaharu Nakayama, Satoshi Yasuda

**Supplemental Figure 1. Deep learning model architecture for ECG-based AS detection**

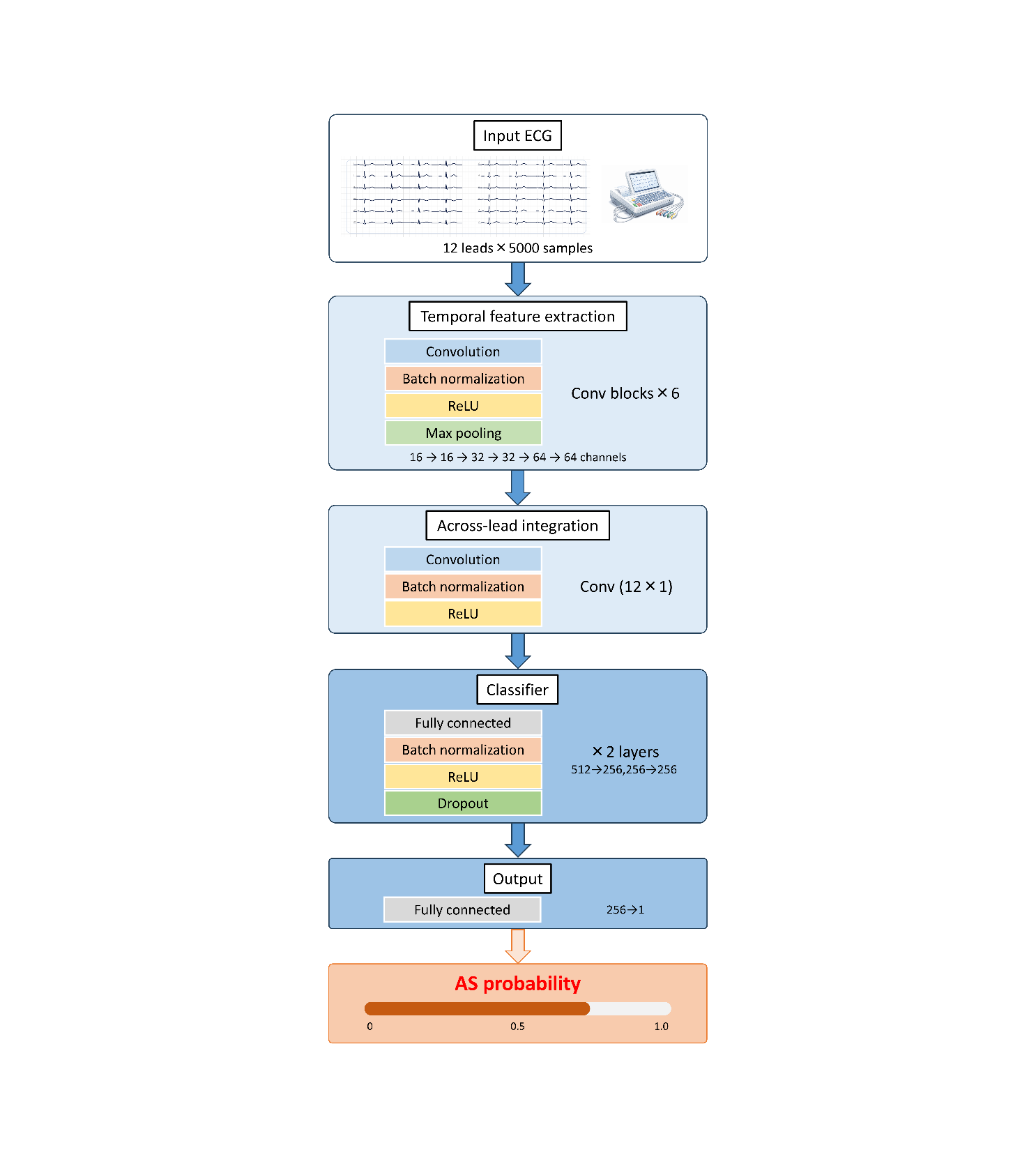

Schematic of the convolutional neural network architecture. Input consists of 12-lead ECG signals (12 leads × 5,000 samples). Temporal features are extracted through six convolutional blocks (Conv-BN-ReLU-MaxPool), followed by across-lead integration via a 12×1 convolution. Two fully connected layers (512→256→256) with batch normalization, ReLU activation, and dropout precede the output layer, which produces a scalar AS probability via sigmoid activation.

Abbreviations: ECG, electrocardiogram; AS, aortic stenosis; Conv, convolution; BN, batch normalization; ReLU, rectified linear unit; MaxPool, max pooling

**Supplemental Figure 2. Identification of R and S wave voltage**

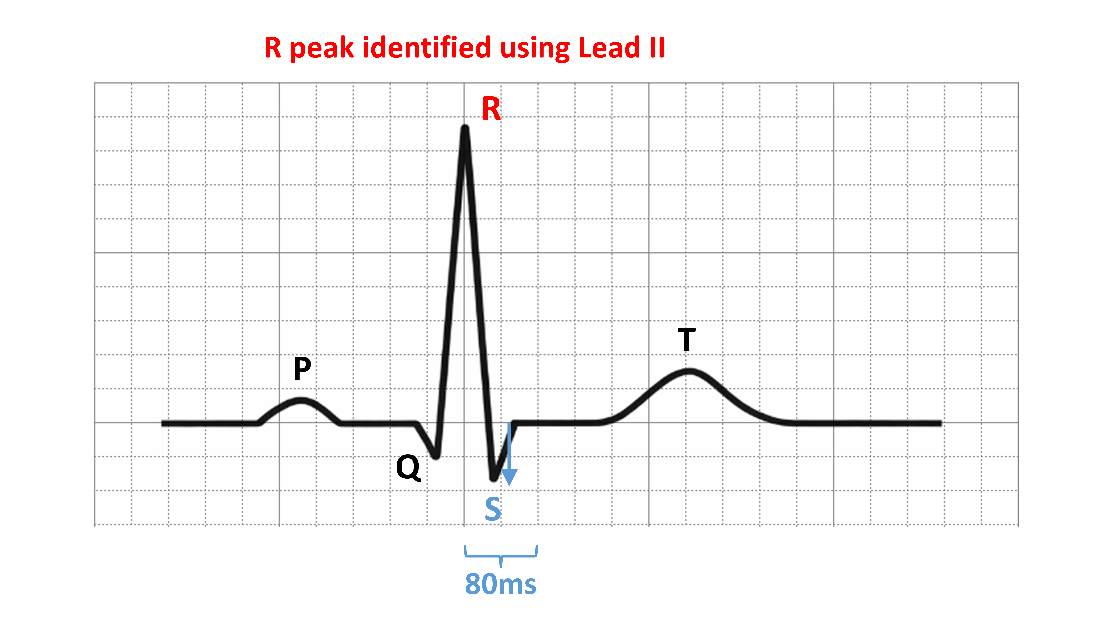

Schematic illustrating the method for identifying R and S wave voltages used to calculate Sokolow-Lyon and Cornell voltage criteria. The R peak was first identified in lead II, and the S wave nadir was defined as the minimum voltage within 80 ms following the R peak in the target lead.

**Supplemental Table 1. AI model discrimination across institutions and subgroups**

| Subgroup | n | AS (n) | AUC (95% CI) |
| --- | --- | --- | --- |
| Institution |  |  |  |
| Kyushu University | 1,687 | 127 | 0.866 (0.838–0.892) |
| Saga University | 1,378 | 61 | 0.859 (0.816–0.902) |
| University of Tokyo | 1,476 | 46 | 0.851 (0.793–0.901) |
| Ehime University | 907 | 56 | 0.845 (0.787–0.896) |
| Osaka Metropolitan University | 1,055 | 115 | 0.830 (0.790–0.869) |
| Kumamoto University | 1,241 | 82 | 0.816 (0.768–0.862) |
| Sex |  |  |  |
| Male | 4,730 | 159 | 0.852 (0.822–0.880) |
| Female | 3,014 | 328 | 0.845 (0.823–0.866) |
| Age group |  |  |  |
| <80 years | 6,184 | 175 | 0.820 (0.790–0.850) |
| ≥80 years | 1,560 | 312 | 0.829 (0.806–0.853) |

Discrimination performance of the AI-ECG model across institutions, sex, and age subgroups in the test cohort, evaluated by AUC with 95% bootstrap confidence intervals.

Abbreviations: AI, artificial intelligence; AS, aortic stenosis; AUC, area under the curve; CI, confidence interval

**Supplemental Table 2. Screening performance at different probability thresholds**

| Threshold | Sensitivity (%) | Specificity (%) |
| --- | --- | --- |
| 0.5 | 20.5 | 98.3 |
| 0.2 | 63.9 | 86.1 |
| 0.1 | 81.7 | 72.7 |
| 0.05 | 90.6 | 56.0 |

The probability threshold of 0.1 was selected based on the validation cohort to maximize sensitivity while maintaining acceptable specificity for screening purposes.

**Supplemental Figure 3. Predictive performance of conventional LVH indices**

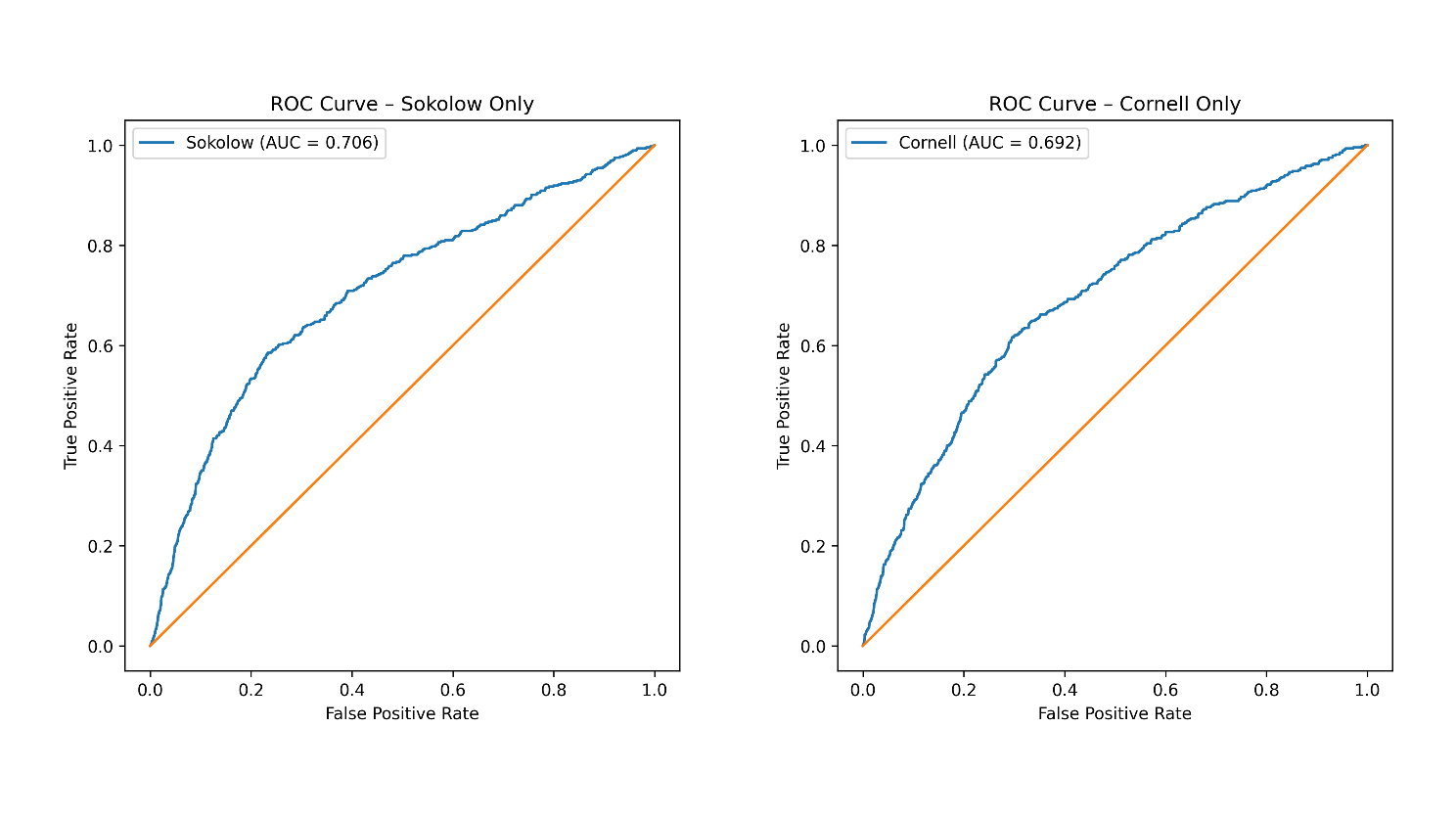

ROC curves for Sokolow-Lyon voltage criterion alone (AUC = 0.706, left) and Cornell voltage criterion alone (AUC = 0.692, right) in the test cohort.

Abbreviations: LVH, left ventricular hypertrophy; ROC, receiver operating characteristic; AUC, area under the curve
